# Attrition and associated factors among patients on chronic antihypertensive therapy at Mulago hospital, Uganda: A mixed method study

**DOI:** 10.1101/2025.06.27.25330319

**Authors:** Nathan Ntenkaire, Mark Kaddu Mukasa, Patience Muwanguzi, Mikka Brian, Sandra Lunkuse, Julius Mubiru, Maxwell Okwero, Beatrice Basuuta, Douglas Bulafu, Joan N Kalyango

**Affiliations:** Clinical Epidemiology Unit, College of Health Sciences, Makerere University, Kampala, Uganda; Infectious Diseases Institute, College of Health Sciences, Makerere University, Kampala, Uganda; Department of Internal Medicine, School of Medicine, College of Health Sciences, Makerere University, Kampala, Uganda; Department of Nursing, School of Health Sciences, College of Health Sciences, Makerere University, Kampala, Uganda; The Medical Research Council/Uganda Virus Research Institute, and LSHTM research unit, Entebbe, Uganda; Department of Disease Control and Environmental Health, School of Public Health, College of Health Sciences, Makerere University, Kampala, Uganda; Department of Pharmacy, School of Health Sciences, College of Health Sciences, Makerere University, Kampala, Uganda

**Keywords:** Attrition, reasons for loss to follow up, antihypertensive therapy, hypertensive patients, Mulago hospital, Uganda

## Abstract

**Background:** Attrition among patients on chronic antihypertensive therapy is a significant problem that can lead to serious health consequences, including uncontrolled blood pressure. Several factors underlie attrition, so healthcare providers must address them to prevent treatment discontinuation and ensure optimal outcomes. Attrition and associated factors was assessed among patients on chronic antihypertensive therapy at Mulago hospital between January 2020 and December 2022.

**Methods:** A sequential explanatory mixed-methods design. The quantitative study was a retrospective cohort study design using files of 1215 hypertensive patients. The qualitative study employed an explanatory descriptive design among 16 patients. A data abstraction tool and an interview guide were used for data collection. Attrition was defined as patients lost to follow-up after missing ≥2 consecutive clinic appointments. Extended Cox regression was used to determine the factors associated with time to attrition at 5% level of significance and qualitative data analysis employed a thematic analysis codebook.

**Results:** The attrition rate was 56.8% (95% confidence interval (CI) 54.0-59.7) with most patients getting lost to follow-up in 2020 (64.9%) and fewest in 2021 (54.7%). Age (hazard ratio (HR)=0.904, 95% CI 0.877-0.932), female Sex (HR=0.713, 95% CI 0.602-0.845), residence outside Kampala (Capital City) (HR=1.311, 95%CI 1.121-1.533), last visit systolic blood pressure (HR=1.013, 95% CI 1.008-1.017), and last visit diastolic blood pressure (HR=0.957,95% CI 0.925-0.990) were associated with time to attrition. Loss to follow-up was driven by structural and contextual barriers, health system challenges, and illness perceptions and health-related limitations.

**Conclusion:** Hypertensive patient attrition rate at Mulago hospital is high, considerably below the Centers for Disease Control and Prevention’s 80% retention target. This calls for innovative retention strategies, and targeted support for high-risk groups like the male patients and those distant from the health facility. Patient-centered approaches addressing structural and health system barriers are essential to improving retention in hypertension care.

## Introduction

Non-communicable diseases (NCDs) are responsible for the majority of deaths worldwide, low and middle-income countries (LMICs) contribute to approximately 75% of all NCD-related fatalities (1). Globally, an estimated 1.28 billion adults (30–79 years old) have hypertension, and the majority (two-thirds) of these live in LMICs (2). In sub-Saharan Africa (SSA), the prevalence of hypertension has risen, with estimates in 2019 reaching 48% (CI: 42–54%) among women and 34% (CI: 29–39%) among men (3). In Uganda, hypertension is estimated to have a prevalence rate of 26.4% in general, with the highest rate of 28.5% occurring in the central region, and the lowest rate of 23.3% in the northern region. (4). Furthermore, the prevalence rates of hypertension in urban and rural areas are estimated to be 28.9% and 25.8%, respectively (5). Only 3.6% of hypertensive patients in Uganda have their blood pressure under control, and hypertension (HTN) and other NCDs account for 33% of all deaths with a 22% probability of dying prematurely from either cardiovascular disease, cancer, chronic respiratory diseases or diabetes (6).

Although HTN cannot be cured, it can be controlled with medication, dietary changes, or a combination of both (7). Patients who consistently engage in medical care at a healthcare facility are considered to be retained in hypertension care. For the long-term management of hypertension and program maintenance, it is critical to have high retention rates. However, in resource-limited settings, 1-year retention rates are often below 50% (8). Retention on antihypertensive therapy is essential for controlling hypertension and lowering the risk of complications related to high blood pressure. Hypertension can lead to various complications, such as cardiovascular disease, kidney disease, cognitive impairment, and eye damage. These complications can cause a range of burdens, including higher healthcare expenses, reduced productivity and quality of life, and an increased risk of disability and premature death (9).

Attrition rate from antihypertensive therapy varies in different studies. Studies done in Cambodia and India reported attrition rates ranging from 9.2% to 61.5% (10, 11). The factors associated with attrition include patient-related factors such as sex, smoking, age, body mass index (BMI). illiteracy, low income, multi-person household (12, 13). There are also clinical factors such as uncontrolled blood pressure, adverse events, medication regimen complex index, and history of hospitalization (14, 15). In addition, health system factors such as stock out, quality of health services, physician-patient relationship, health education and availability of medication have been associated with attrition (16). With the rising prevalence of chronic diseases, health systems in Africa are struggling to maintain continuity of care (17), yet no research has been conducted in Uganda to determine the attrition rate of patients with hypertension and it is hypothesized that rates of attrition are high and its predictors are varied. Additionally, patients view on the reason for loss to follow up from hypertension care is less known, Therefore, this study determined attrition rate and associated factors among patients on chronic antihypertensive therapy at Mulago hospital and explored reasons for loss to follow-up from the perspective of patients identified as lost to follow-up.

## Materials and methods

### Study setting

The study was conducted at the hypertension clinic in the medical outpatients’ department of Mulago Hospital, which is situated in Kampala, Uganda’s capital city. The hospital serves as the primary teaching facility for the College of Health Sciences at Makerere University. It offers comprehensive healthcare services across numerous medical and surgical subspecialties, including dentistry, emergency medicine, pediatrics, and intensive. The clinic is open on Mondays, except on public holidays, and is staffed by specialist physicians, including a cardiologist, nurses, and records officers as well as laboratory and pharmacy support. Approximately 438 patients are initiated on antihypertensive treatment (AHT) each year at this clinic.

### Study design and population

A sequential explanatory mixed methods design, consisting of two distinct phases by Tashakkori and Creswell (18) was adopted. The quantitative study phase was a retrospective cohort, and the qualitative study phase was a descriptive explanatory design. For the quantitative study, files of patients on chronic antihypertensive therapy who were registered at the Mulago hypertension clinic between January 2020 and December 2022 were included. Patients’ files missing data on more than 30% of the studied variables were excluded. Patients on chronic antihypertensive therapy who were registered at the Mulago hospital between January 2020 and December 2022, who were confirmed as lost to follow up and consented to participate were included in the qualitative study. Patients who were transferred to other health facilities and those who did not understand English or Luganda (the most commonly spoken local language in the area) were excluded from participation in the interviews.

### Sample size and Sampling procedure

Sample size for the objective of determining the attrition rate was calculated using the Kish (1965) formula for determining sample size (19). Considering the study done by Meena and colleagues that reported an attrition rate among hypertensive patients of 9.2% (20), and a sampling error of 5%. A sample size of 397 was required.

To determine the factors associated with time to attrition, the sample size was estimated using the formula for survival data sample size calculation. In this study, 5% level of significance, a two-sided test in analysis, and 80% power for estimated sample size were assumed. Considering what Given and colleagues reported among patients on antihypertensive treatment where 67.9% of the 53 patients aged 41-52 and 79.6% of 54 patients aged 53-65 were retained in care respectively. (21).

The estimated sample size was based on the number of events.

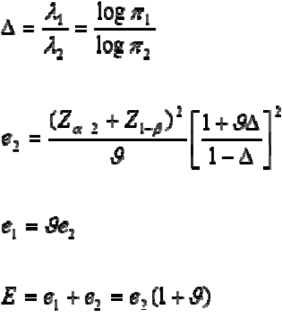

Where,

π1 = Proportion surviving (retained on treatment) in those aged 41-52 (67.9%)

π2 = Proportion surviving (retained on treatment) in those 53-65 (79.6%)

Zα/2= Standard normal value corresponding to a 5% level of significance (1.96)

Z1-β = Standard normal value corresponding to 80% power of study (0.84)

ϑ = Ratio of those aged 53-65 to those aged 41-52 (54/53)

e1= Expected events in those aged 41-52

e2=Expected events in those aged 53-65

E= Number of events

Substituting in the above formulae, 238 events were gotten. Then the number of events were substituted in the formula below.

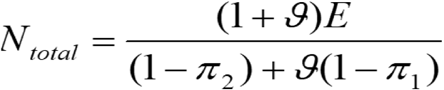

Therefore, the required number of patient files was 905, and after accounting for missing data and assuming a 10% missing data, a sample size of 1006 patient files was needed.

Consecutive sampling technique was used to select patient files of hypertensive patients who were registered at Mulago hospital. Participants for the in-depth phone interviews were selected using purposive sampling with maximum variation, considering factors such as age, sex, residence, presence of comorbidities, and adverse events, from among those identified as lost to follow-up. The final number of interviewees was determined at the point of data saturation, when no new information emerged and sufficient depth on the topic had been achieved.

### Variables

The dependent variable was time to attrition and the independent variables were patient-related factors: age, sex, residence and occupation, alcohol use,smoking status, herbal medicine use, and marital status.

Clinical-related factors: Baseline SBP and DBP, presence of comorbidities, history of hospitalization, number of antihypertensive drugs, side effects, hypertension drug regimen, and last visit SBP and DBP. Health system-related factor: medicine stock out.

### Data collection

Secondary data obtained in the hospital for patients’ clinical monitoring and evaluation was used in this study and it was accessed between 07^th^ July 2023 to 20^th^ November 2023.

The primary source of data was the patient files kept at the hypertension clinic and the pharmacy register to complement the information. The data abstraction tool was pretested on 10 patient files and standardized before being used for actual data collection and these 10 patient files used for pretesting were not part of the final patient files considered. Demographic, clinical and health system data was collected by two trained registered clinic nurses. The Principal Investigator (PI) regularly double-checked the filled data abstraction tool by trained registered clinic nurses against the patient files to ensure that the data collected was accurate and free of errors.

For the qualitative component of the study, two trained and registered clinic nurses contacted patients who had been confirmed as lost to follow-up (LTFU) and guided them through the informed consent process. The participants were recruited from 20th January 2024 to 28th January 2024. Those who provided consent were interviewed by phone to explore the reasons for their loss to follow-up. The interviews were conducted using an interview guide developed in both English and Luganda, based on participant language preference. Interviews were audio recorded with prior permission from the participants and were held in the records office, in the presence of the Principal Investigator (PI). Each interview lasted a maximum of 30 minutes. To minimize loss of information, field notes were also taken in notebooks during each session. Data collection continued until saturation was achieved, which occurred after 16 interviews. To ensure the credibility and dependability of the findings, patient verification and peer-review quality control practices were employed. Transcripts were not returned to participants for feedback due to nature of interviews.

### Data management

The collected quantitative data was entered into EpiData Manager version 4.7.0, where it was verified for accuracy, consistency, and completeness. It was then cleaned and edited before being analyzed using Stata version 15.0. For qualitative data, audio recordings in English and Luganda were transcribed verbatim and translated into English text before analysis.

Both qualitative and quantitative data were securely stored on a password-protected computer to ensure data confidentiality.

### Data analysis

Descriptive analysis where measures of central tendency and dispersion (mean, standard deviation, median and interquartile ranges (IQR)) were computed for numerical variables. For categorical variables, frequencies and percentages were computed, and tables and graphs were used to visualize the analyzed data. A proportion was obtained to determine the percentage of patients that experienced attrition by dividing the number of hypertensive patients who were lost to follow-up by the study sample size.

The probabilities of patients staying in care at different intervals of the follow-up period were determined using the Kaplan-Meier method, and the log-rank test was used to determine the significance of observed differences between groups. The variables were assessed for fulfillment of Cox proportional hazards assumptions using graphical approaches, test residuals and also extend the model to include time varying covariates-During this assessment, we found that one of the variables violated this assumption hence the Extended Cox regression model was used in all analyses that included this variable to determine the factors associated with time to attrition.

Variables with p-values of ≤ 0.2 at bivariate analysis and those known to be associated with attrition from literature were considered for multivariate analysis. A chunk test was used to compare the reduced and full model and therefore assess for interaction. Confounding was then assessed where a variable was considered a confounder if the change in the hazard ratio (HR) was > 10%. Hazard ratios (HR), p-values and the 95% confidence intervals were reported.

Qualitative data analysis involved the utilization of a thematic analysis codebook, applying the 6 phases inherent in the thematic analysis (TA) approach (22).The data was transcribed, translated in English, coded, and synthesized using Open Code version 4.02 to yield notable themes (23). Three trained data coders independently coded the transcripts to enhance reliability and reduce individual coder bias. A thematic analysis codebook was applied, and themes were inductively derived from the data. Discrepancies in coding were resolved through discussion and consensus among the coders. While rigorous analytical procedures were followed, participants did not provide feedback on the findings.

### Ethical considerations

Permission to conduct the study was granted by the Clinical Epidemiology Unit (CEU). Ethical approval was obtained from the Institutional Review Board (IRB) through the School of Medicine Research and Ethics Committee (SOMREC) of Makerere University College of Health Sciences (Mak-SOMREC-2023-584). Additionally, SOMREC approved a waiver of consent to allow the use of patient records. Written informed consent was obtained from participants confirmed to be lost to follow-up who took part in the qualitative phase of the study. The principal investigator and the two trained registered clinic nurses were involved in reviewing patient files and information obtained from the patient files was not disclosed to any third person. Anonymity for all patients was considered and identification variables such as names were omitted during data extraction.

## Results

Among the 1,278 patients registered at the Mulago Hypertension Clinic (2020–2022), 63 were excluded due to missing more than 30% of study data. A total of 1215 patient files meeting the eligibility criteria were selected for the quantitative study. Of these, 20 participants were selected to take part in the in-depth phone interviews (Fig 1).

**Fig 1.**
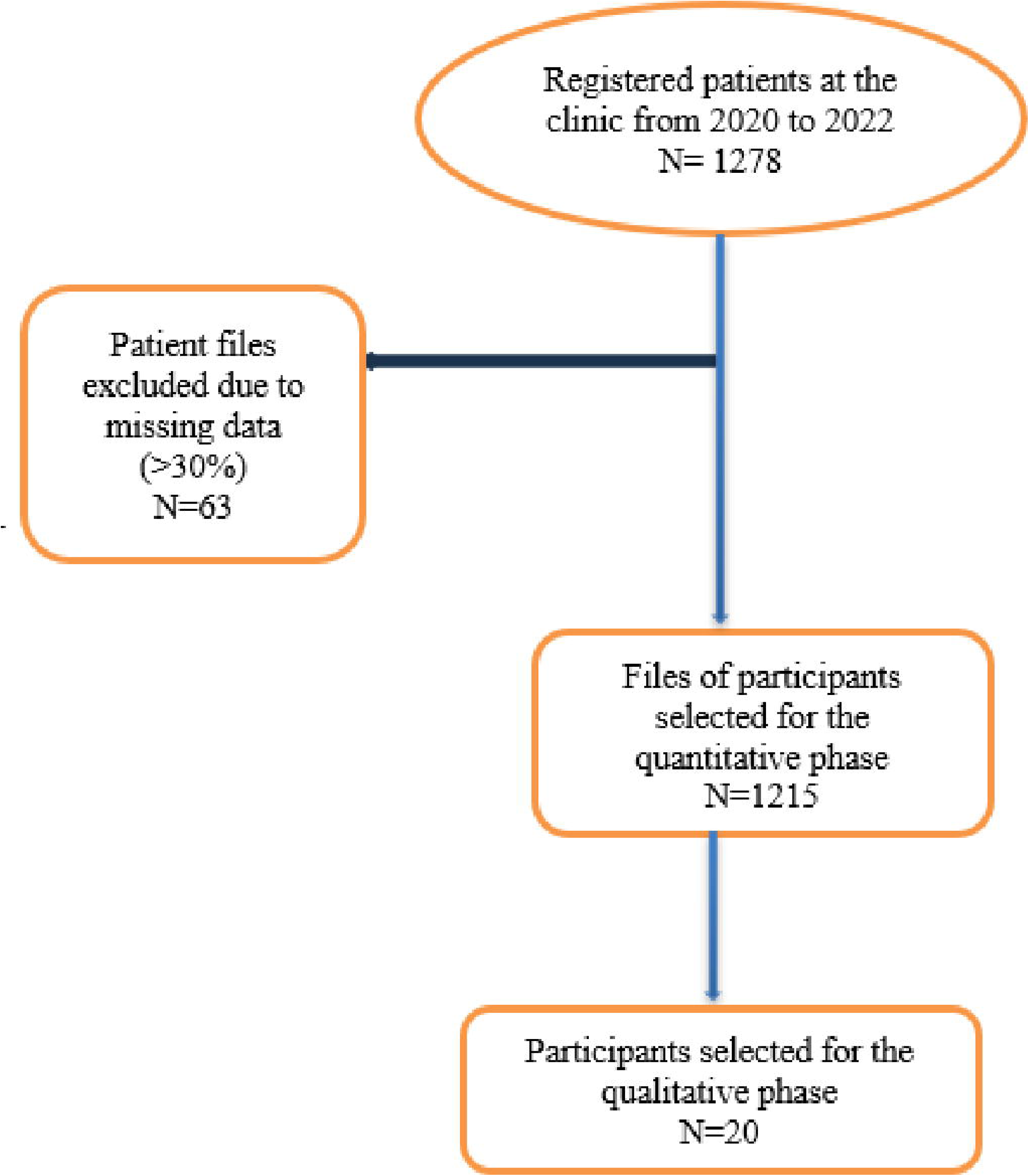

### Characteristics of patients registered for anti-hypertensive therapy at the Mulago hospital from January 2020 to December 2022

More than half of the patients resided outside Kampala (58.8%, n= 714) and majority were female (76.1%, n= 924), with mean age of 56.5 years (SD 13.9). The majority did not smoke (99.4%, n=1208), while 98.2% had no history of alcohol use (98.2%, n=1193). About half had a comorbidity (50.8%, n=617), while 92.8% (n=1128) had never been hospitalized and 58.5%(n=711) had experienced side effects. The median baseline systolic blood pressure (1^st^, 3^rd^ quartile) was 150 (138, 168) and the median baseline diastolic blood pressure was 87 (77, 97).

Regarding the drug regimen, the majority were on combination therapy (86.3%, n=1048) and the least on diuretics (0.08%, n=10). Most of the patients (59.6%, n= 724) had prescriptions of ≤ 2 antihypertensive drugs. Medicine stock outs were reported in 80.3% (n=922) of the patients during at least one of the visits.

A small part of the patients used herbal medicine (1.0%, n= 12). On the last clinic visit, the median systolic blood pressure was 145 (132, 159), and the median diastolic blood pressure was 83 (75, 92). Business was the most common occupation among the patients (29.1%, n=188) followed by peasants (24.6%, n=159) (Table 1).

**Table 1.**
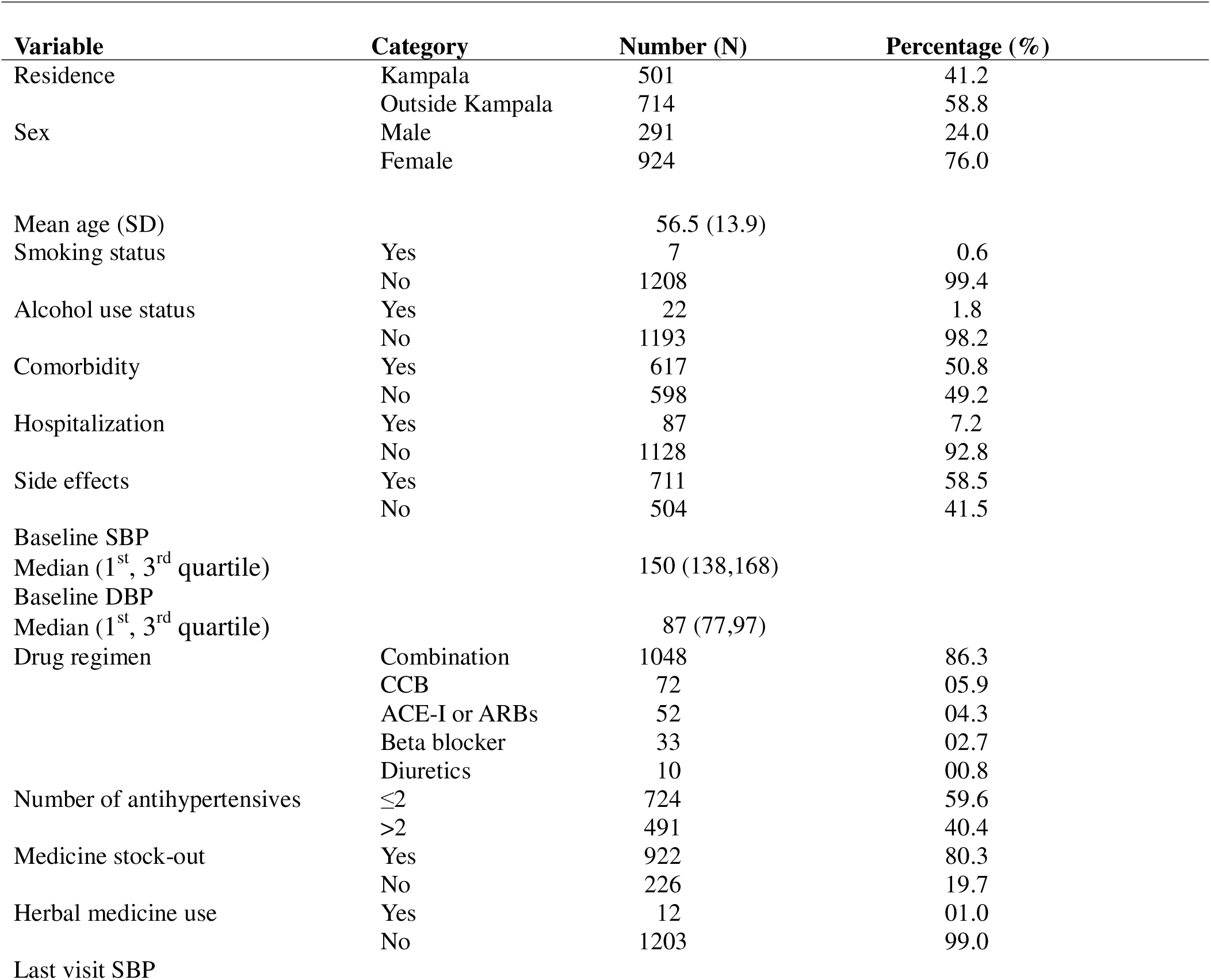
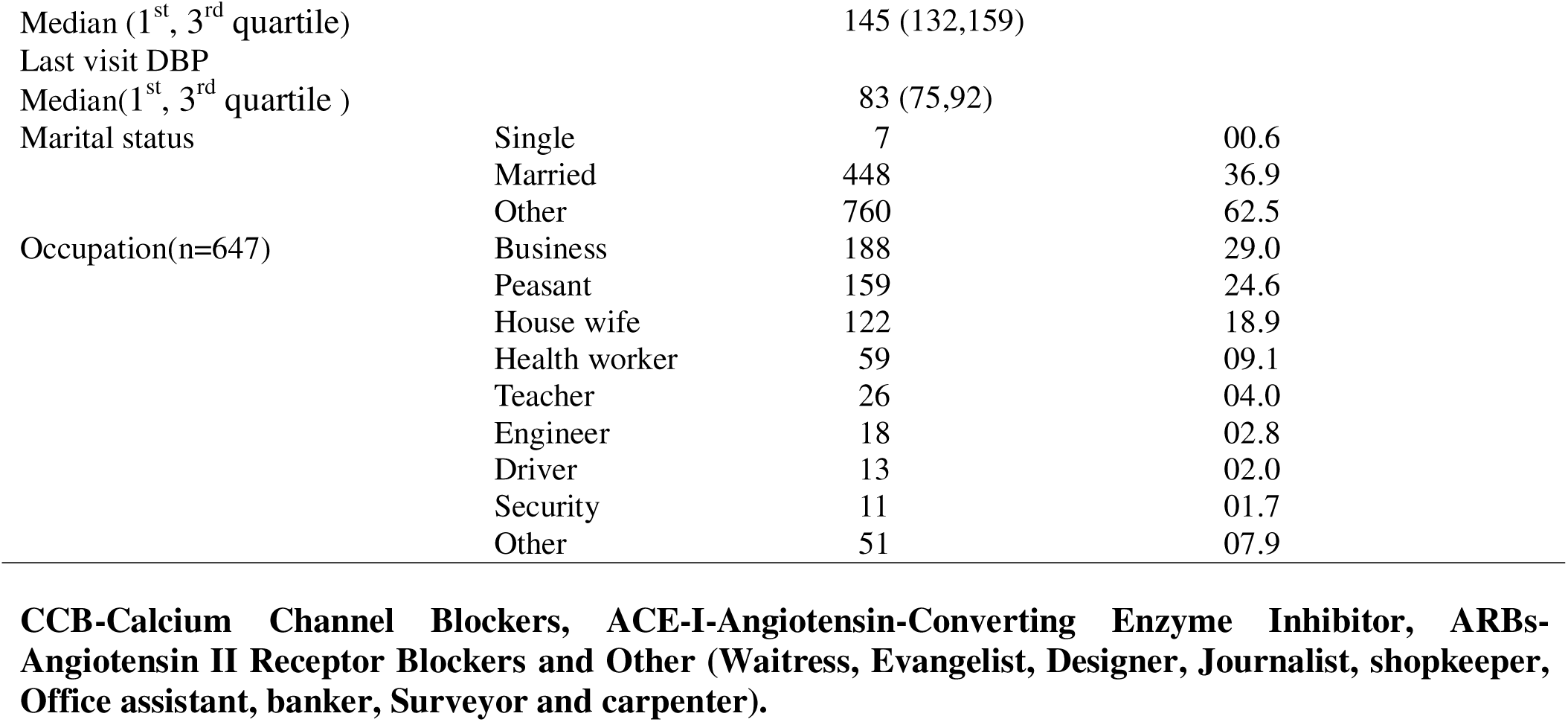
Characteristics of patients registered for antihypertensive therapy at Mulago hospital, Jan 2020-Dec-2022 (n=1215)

### Attrition rate among patients on chronic antihypertensive therapy at Mulago hospital initiated on AHT between Jan 2020 and Dec 2022

The overall attrition rate was 56.8 % (95% CI, 54.0-59.6). Patients registered in 2020 had the highest attrition rate at 64.9%, followed by 2022 at 55.0 %, and 2021 had a relatively reduced attrition rate of 54.7% When considering sex, male patients had a greater attrition rate (64.6%) than females (54.3%). Furthermore, patients residing outside Kampala had a relatively higher attrition rate (59.8%,) than those dwelling within Kampala, where the attrition rate was 52.6 % (Fig 2)

**Fig 2.**
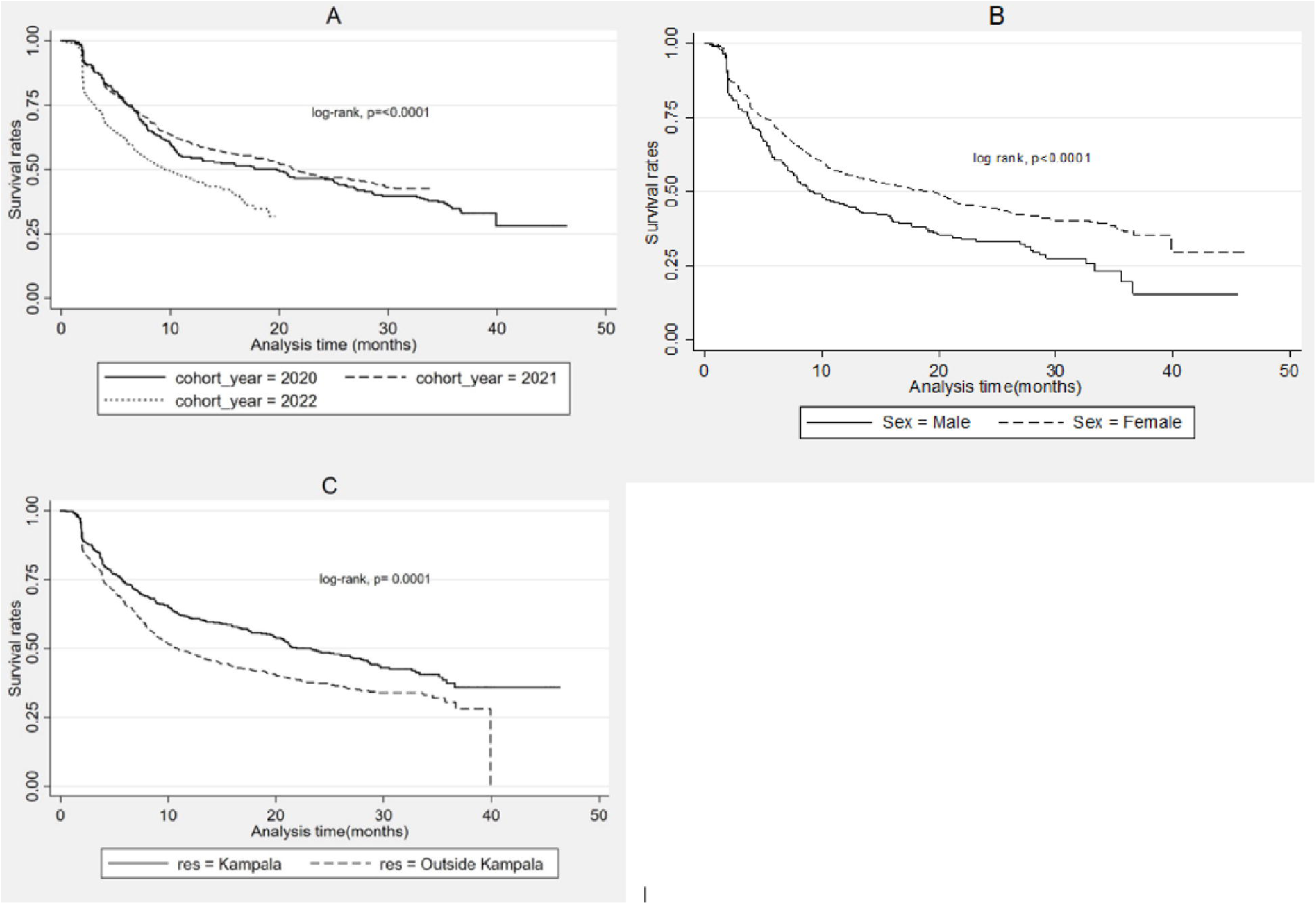

### Factors associated with time to attrition among patients on chronic antihypertensive therapy at Mulago Hospital between January 2020 and December 2022

In bivariate analysis factors such as residence, sex, smoking, age, side effects, baseline systolic blood pressure (SBP), drug regimen, SBP on the last visit, and diastolic blood pressure (DBP) on the last visit were selected for multivariate analysis (Tables 2 and 3). In multivariate analysis factors associated with attrition were age (HR=0.904, 95% CI 0.877-0.932), residence: Outside Kampala (HR=1.311,95%CI 1.121-1.533), Sex: Female (HR=0.713, 95% CI 0.602-0.845), SBP on last visit (HR=1.013,95% CI 1.008-1.017) and DBP on last visit (HR=0.957,95% CI 0.925-0.990). DBP on the last visit was a time-varying covariate with p=0.006 (Table 4).

**Table 2.** Bivariate analysis for patient related factors associated with time to attrition among 1215 patients on chronic antihypertensive therapy at Mulago hospital, Jan 2020-Dec 2022.

| Variable | HR (95% CI) | p-value |
| --- | --- | --- |
| **Residence** |  |  |
| Kampala | Ref |  |
| Outside Kampala | 1.374 (1.176-1.606) | <0.001 |
| **Sex** |  |  |
| Male | Ref |  |
| Female | 0.706 (0.597-0.835) | <0.001 |
| **Age** | 0.999 (0.993-1.004) | 0.637 |
| **Smoking status** |  |  |
| Yes | Ref |  |
| No | 0.682 (0.305-1.523) | 0.351 |
| **Alcohol use** |  |  |
| Yes | Ref |  |
| No | 0.896 (0.528-1.521) | 0.685 |
| **Occupation** |  |  |
| Business | Ref |  |
| Peasant | 0.650 (0.551-1.042) | 0.245 |
| House wife | 0.980 (0.678-3.050) | 0.201 |
| Health worker | 0.450 (0.310- 2.890) | 0.254 |
| Teacher | 0.720 (0.525-1.150) | 0.325 |
| Engineer | 0.691 (0.615-1.810) | 0.980 |
| Driver | 1.011 (0.434-1.431) | 0.421 |
| Security | 1.510 (0.781-3.100) | 0.267 |
| Other | 0.890 (0.343-2.194) | 0.723 |

**HR= hazard ratio, CI=confidence interval**

**Table 3.** Bivariate analysis for clinical and health facility related factors associated with time to attrition among 1215 patients on chronic antihypertensive therapy at Mulago hospital, Jan 2020-Dec 2022.

|  |  |  | tvc |  |
| --- | --- | --- | --- | --- |
| Variable | HR (95% CI) | p-value | HR (95% CI) | p-value |
| Comorbidity |  |  |  |  |
| Yes | Ref |  |  |  |
| No | 1.149 (0.989-1.334) | 1.149 |  |  |
| Hospitalization |  |  |  |  |
| Yes | Ref |  |  |  |
| No | 0.798 (0.533-1.193) | 0.271 |  |  |
| Side effects |  |  |  |  |
| Yes | Ref |  |  |  |
| No | 1.134 (0.974-1.320) | 0.107 |  |  |
| Baseline SBP | 1.002 (0.999-1.005) | 0.178 |  |  |
| Baseline DBP | 1.000 (0.995-1.005) | 0.934 |  |  |
| Drug regimen |  |  |  |  |
| CCB | Ref |  |  |  |
| Combination | 0.769 (0.568-1.042) | 0.090 |  |  |
| ACE-1 or ARBs | 0.692 (0.427-1.122) | 0.135 |  |  |
| Beta blockers | 1.381 (0.852-2.239) | 0.190 |  |  |
| Diuretics | 0.825 (0.352-1.934) | 0.658 |  |  |
| **Number of antihypertensives** |  |  |  |  |
| ≤2 | Ref |  |  |  |
| ≥2 | 0.925 (0.794-1.077) | 0.315 |  |  |
| **Medicine stock out** |  |  |  |  |
| Yes | Ref |  |  |  |
| No | 0.878 (0.714-1.080) | 0.217 |  |  |
| **Herbal use** |  |  |  |  |
| Yes | Ref |  |  |  |
| No | 1.646 (0.683-3.969) | 0.267 |  |  |
| **Last visit SBP** | 1.009 (1.006-1.013) | <0.001 |  |  |
| **Last visit DBP** | 1.015 (1.006-1.023) | 0.001 | 0.998 (0.998 -0.999) | <0.001 |

HR= hazard ratio, CI=confidence interval, tv= time varying covariates.

**Table 4:** Multivariate analysis for predictors of time to attrition among 1215 patients on chronic antihypertensive therapy at Mulago hospital, Jan 2020-Dec 2022.

|  |  |  | tvc |  |
| --- | --- | --- | --- | --- |
| Variable | HR (95% CI) | p-value | HR (95% CI) | P-value |
| Residence |  |  |  |  |
| Kampala | Ref |  |  |  |
| Outside Kampala | 1.311 (1.121- 1.533) | 0.001 |  |  |
| Sex |  |  |  |  |
| Male | Ref |  |  |  |
| Female | 0.713 (0.602- 0.845) | <0.001 |  |  |
| SBP on last visit | 1.013 (1.008-1.017) | <0.001 |  |  |
| **DBP on last visit** | 0.957 (0.925-0.990) | 0.011 | 0.9989(0.9981-0.9997) | 0.006 |
| **Age** | 0.904(0.877-0.932) | <0.001 |  |  |

**HR= hazard ratio, CI=confidence interval, tv= time varying covariates.**

### Description of the participants that participated in the phone interviews

There were 20 patients out of the 690 found to be lost to follow up that were selected for the interviews, however only 16 participated in the in-depth phone interviews. The mean age of the participants was 50 years (SD 11.99). Nine (56.3%) of the participants were female and 11 (68.8%) of the participants were residing outside Kampala. Five (31.5%) of the participants had a comorbidity, 11(68.8%) of the participants had experienced drug related side effects and 2 (12.5%) of participants reported to have been hospitalized in the past (Table 5).

**Table 5:** Description of the 16 participants that took part in the phone interview.

| **Characteristics** | **Category** | **n (%)** |
| --- | --- | --- |
| Sex | Male | 7 (43.8) |
|  | Female | 9 (56.2) |
| Age |  |  |
| Mean (SD) |  | 50 (12.0) |
| Residence | Kampala | 5 (31.3) |
|  | Outside Kampala | 11 (68.7) |
| Presence of comorbidity | Yes | 5 (31.3) |
|  | No | 11 (68.7) |
| Presence of side effects | Yes | 11 (68.7) |
|  | No | 5 (31.3) |
| Hospitalization | Yes | 2 (12.5) |
|  | No | 14 (87.5) |

### Underlying reasons for loss to follow up among patients on chronic antihypertensive therapy at Mulago hospital hypertension clinic

Participants reported a variety of factors contributing to loss to follow-up (LTFU) from hypertension care. These were grouped into three major themes: (1) Structural and Contextual Barriers, (2) Health System Barriers, and (3) Illness Perceptions and Health-Related Limitations. Each theme comprised of multiple subthemes (Fig 3).

**Fig 3.**
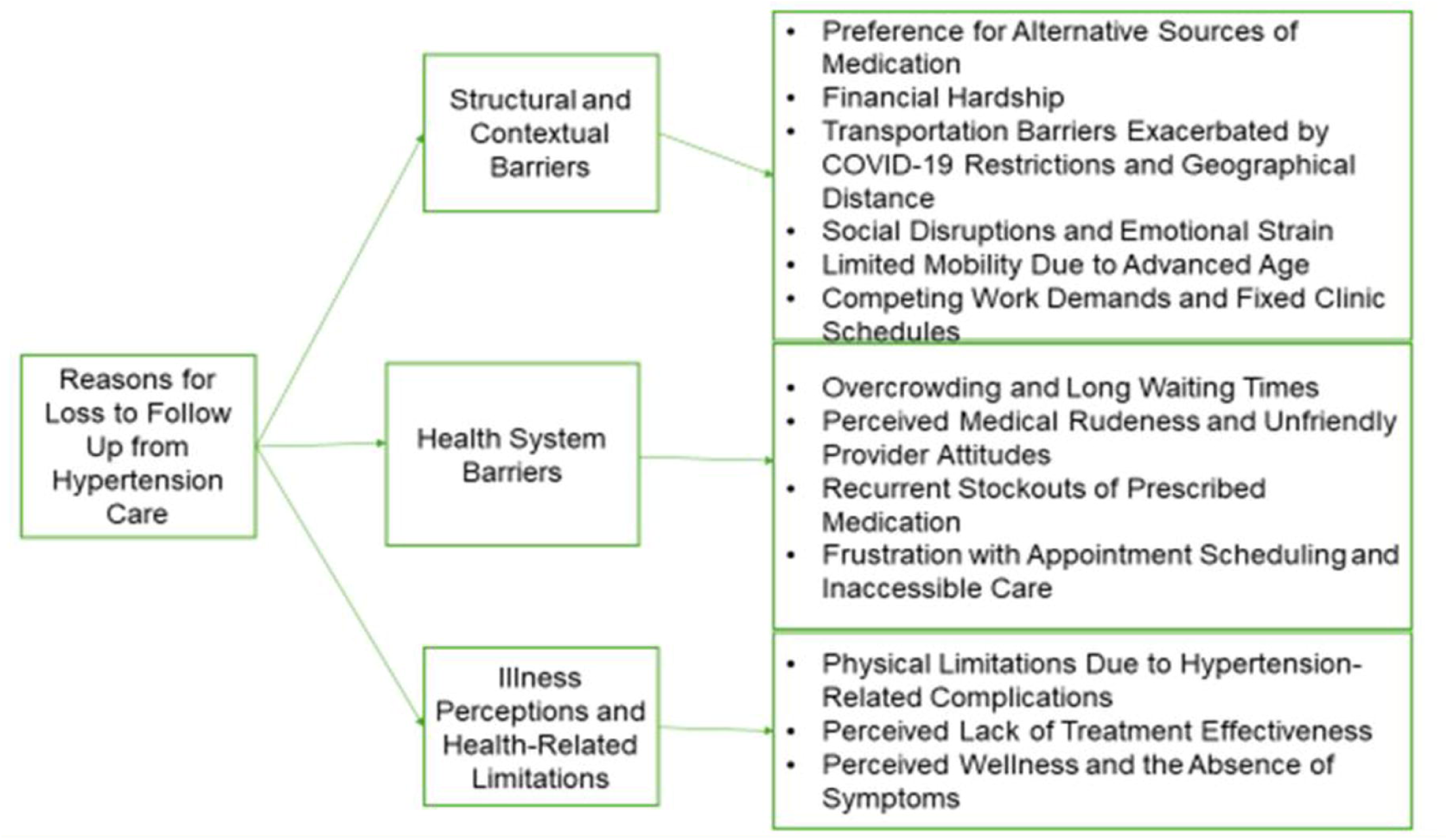

#### Theme 1: Structural and Contextual Barriers

##### Preference for Alternative Source of Medication

Participants obtained medication from nearby private clinics and pharmacies, with some preferring herbal remedies as alternatives to conventional medicine

‘’I’m also a health work, most of the time I get my drugs from somewhere else, I buy from a pharmacy nearby home…….” (**Female, lost to follow-up**), “…….my sibling at Najjanakumbi gave me some herbal medicine, that I use………” (**Female, lost to follow-up**).

##### Financial Hardship

Economic hardship was a key deterrent to continued care as participants could not afford transportation to the clinic.

*“……. I’ve been having a challenge of lack of money; I’ll harvest some maize to see if I can generate funds to come …….”* (**Male, lost to follow-up**).

##### Transportation Barriers Exacerbated by COVID-19 Restrictions and Geographical Distance

Participants reported that pandemic-related restrictions and long travel distances to the clinic affected their ability to attend appointments.

***“……****the problem is COVID-19 came in and destabilized movement, even public means we use was stopped…….”* (**Female, loss to follow up**), “……. *I met some people and they told me to go to Kasese hospital for treatment because the distance to Mulago was far*” (**Male, lost to follow-up**).

##### Social Disruptions and Emotional Strain

Participants reported bereavement, prolonged travel for burial ceremonies, and lack of support or caretakers as key reasons for missing hypertension clinic follow-up visits.

*“……I had some problems; I lost my relative and I traveled for burial and took long to come back”* (**Female, loss to follow-up**), *“……I don’t have a caretaker, it is me who takes care of myself, my children who would take care of me are not around…”* (**Female, lost to follow-up**).

##### Limited Mobility Due to Advanced Age

Elderly participants reported mobility limitations as a reason for their loss to follow up.

“ being an elderly person and weak, I was tired and decided to just sit home and leave alone with going to the clinic……” (**Female, loss to follow-up**).

##### Competing Work Demands and fixed clinic schedules

Work-related obligations conflicted with non-flexible clinic schedules, led to loss to follow up.

*“… work is too much at the specialized hospital where I work, a few times I visited clinic people at work complained thinking I had gone to work somewhere else…”* (**Female, lost to follow-up**).

#### Theme 2: Health System Barriers

##### Overcrowding and Long Waiting Times

Overcrowding and waiting for long before served at the clinic discouraged participants from returning for follow-up visits.

*“……. sometimes you reach at the clinic and you are made to stay in the queue for so long”* (**Female, loss to follow-up**), ***“…*** *I came to the clinic, there were very many patients and we would spend a lot of time*

*there…….”* (**Female, loss to follow-up**).

##### Perceived Medical Rudeness and Unfriendly Provider Attitudes

Negative interactions with healthcare providers led to dissatisfaction and disengagement.

*“ most of the doctors are rude, you ask them a question and he is rude and doctors are not willing to help….”* (**Female, loss to follow-up**).

##### Recurrent Stockouts of Prescribed Medication

Inconsistent availability of medication at the clinic pharmacy discouraged patients from returning.

*“……even when you get transport money, you don’t find medicine….”* (**Female, loss to follow-up**)

##### Frustration with Appointment Scheduling and Inaccessible Care

Appointment frustration was a major reason participant missed follow-ups, as scheduling and securing timely visits were challenging.

*“……I was told to go see some doctor but I couldn’t find him for three weeks I came……”* (**Male, loss to follow-up**).

#### Theme 3: Illness Perceptions and Health-Related Limitations

##### Physical Limitations Due to Hypertension-Related Complications

Physical impairments resulting from complications of hypertension also impeded attendance of clinic visits.

*“……I stopped coming because I had a stroke and so it was hard for me to move….”* (**Male, loss to follow). Perceived Lack of Treatment Effectiveness**

Some participants lost confidence in their treatment regimen, expressing doubts about its effectiveness.

*“……. I would not get any change after getting the medication……”* (**Female, loss to follow-up**)

##### Perceived Wellness and the Absence of Symptoms

Several participants discontinued follow-up because they felt physically well and did not perceive a need for ongoing care.

*“…. if I’m feeling well, is there need to come back to the clinic….”* (**Female, loss to follow-up**).

## Discussion

The study found an attrition rate of 56.8% implying that approximately 57 patients on chronic antihypertensive therapy at Mulago hospital were lost to follow up per 100-person month of follow up. This suggests that the retention rate is quite low to achieve the WHO targeted treatment goal of bp <140/90 mmHg in all patients with hypertension These results were consistent with those from the studies by Engelland and colleagues (24) conducted in 4403 hypertensive patients, Ramsay and colleagues (25) conducted among 40 hypertension patients attending a hypertension clinic over 15 months, and Ma and colleagues (11) conducted in China among 520 patients with hypertension which reported 51%, 50% and >30% attrition rate among hypertensive patients respectively.

However, these results were different from the studies by Meena and colleagues (26) conducted on a group of 1036 hypertensive patients who were registered at the NCD clinic in Pratap Nagar, Jodhpur, India and Kassavou and colleagues (27) conducted among 101 hypertensive people in a primary health care which reported a lower attrition rate of 9.2% and 7.92 % respectively among people with hypertension. The difference in the attrition rates could be due to the disparity in the study designs, sample size and study population. Meena, Raghav (26) conducted a longitudinal study while Kassavou, Mirzaei (27) conducted a randomized clinical controlled trial, additionally they used a sample size of 1036 and 102 patients respectively.

The patient’s place of residence was statistically associated with time to attrition among hypertensive patients. Patients who resided outside Kampala were 31.1% more likely to get lost to follow up compared to those who resided in Kampala. This might be due to the fact that individuals residing in Kampala had easy access to the clinic, being in close proximity to it. This explanation aligns with the qualitative findings, where living far away was identified as one of the key reasons for patients being lost to follow-up while

undergoing chronic antihypertensive treatment. Similarly, A. Baldé and colleagues concurred in their findings, indicating that residing more than 5 km away from a healthcare facility was linked to an increased risk of loss to follow-up among individuals living with HIV in Mali, a comparable chronic condition (28).

Patient sex was statistically associated with time to attrition among hypertensive patients. Female patients were 28.7% less likely to be lost to follow up than their male counterpart. These findings are consistent with studies by Degoulet and colleagues (29) conducted among 1346 medical records of hypertensive patients in Paris, and Hernandez and colleagues (30) conducted among 6677 patients with hypertension or diabetes in Cambodia. This could be because women are more likely to seek medical attention than the men. However a study by Given and colleagues (31) found Sex not to be a significant predictor of attrition. This might stem from the fact that Given and colleagues (31) conducted a randomized controlled trial with a small sample size (153) compared to this study.

Age was significantly associated with time to attrition among patients with hypertension. For every unit increase in age of the patient, there was 9.6% decrease in time to attrition. This finding is in agreement with what was reported by Hernandez and colleagues (13) in a study conducted in Cambodia among antihypertensive patients. However, this finding diverges from our qualitative aspect which found advanced age as a reason for loss to follow up. The disparity could be due to the fact that all patients were adults and most elderly. Elderly patients tend to be concerned about their health compared to young individuals who exhibit less interest.

Blood pressure measurements on the last visit date to the clinic were significantly associated with time to attrition. For every unit increase in systolic blood pressure at the last visit to the clinic, there was 1.3% increase in time to attrition. Additionally, for every unit increase in diastolic pressure at the last visit to the clinic, there was 4.3% decrease in time to attrition and at any given time as it increases, the time to attrition decreases by 0.0011%. This could either be due to patients perceived wellness and or lack of improvement while on drugs, which reasons were both highlighted in the qualitative findings. DBP levels change over time due to lifestyle modification, inconsistent adherence to medication and treatment adjustments by health care providers.

Drug regimen was not associated with time to attrition in this study. Although no studies have linked drug regimens to attrition among hypertensive patients, drug regimens have been associated with attrition among 58,115 people living with HIV (PLHIV) in China, a chronic illness similar to hypertension (32). The difference in findings may be attributed to the variation in study populations, as Zhu and colleagues(32) conducted their research among people living with HIV.

The number of antihypertensive drugs was not associated with time to attrition among hypertensive patients. These findings are in agreement with a follow up study conducted by Given and colleagues Given, Given (31) on previously diagnosed 158 patients with hypertension. However a study by Hernandez and colleagues (30) in Cambodia among 6677 patients with hypertension or diabetes revealed number of medications taken to be associated with attrition. The discrepancy between the two studies could be due to the fact that Hernandez and colleagues (30) conducted their study within a larger population of both hypertensive and diabetic patients compared to the current study.

There was no association between history of hospitalization and time to attrition, however a multicenter randomized two-arm controlled trial (ERMIES) conducted by Flaus-Furmaniuk and colleagues in reunion Island among people with type two diabetes, a similar chronic illness to hypertension revealed history of hospitalization to be associated with higher risk of attrition (33). The disparity could be due the fact that Flaus-Furmaniuk and colleagues (33) conducted a randomized controlled trial among a smaller population of 100 type 2 diabetic patients unlike this study which was a retrospective analysis of 1215 hypertensive patient files.

The occurrence of drug related side effects was not associated with time to attrition. Additionally, no studies have reported side effects to be associated with attrition among hypertensive patients. However, Zemarian and colleagues (34) found that drug adverse events were associated with attrition among HIV-infected children in Ethiopia, a chronic illness comparable to hypertension. This variation in findings may be attributed to differences in the study population where Zemarian and colleagues conducted their study among children living with HIV.

Findings from this study found smoking status not to be associated with time to attrition. However a study conducted by Hernandez and colleagues (13) in Cambodia among 6677 patients with hypertension or diabetes found smoking status to be significantly associated with attrition. The discrepancy in the findings could be due to the fact that Hernandez and colleagues (13) conducted their study among a larger number of both hypertensive and diabetic patients unlike this study.

Medicine stock out was not associated with time to attrition. However, a study conducted by Pasquet and colleagues (35) in Abidjan, Côte d’Ivoire among HIV infected patients found drug stock to be associated with interruption from care. The findings from the qualitative aspect are in agreement with the findings from this study as medicine stock-out was highlighted by participants as one of the reasons for loss to follow up.

The qualitative findings reveal a multidimensional interplay of why patients get lost to follow-up, categorized into structural and contextual barriers, health system barriers, and illness perceptions and health-related limitations. Structural and contextual issues such as financial hardship, transportation challenges especially heightened by COVID-19 restrictions and competing work demands illustrate the broader socioeconomic constraints that hinder continuity of care, consistent with findings from prior studies in developed and low-resource settings (36-38). Health system barriers, including long waiting times, perceived provider rudeness, and frequent medication stockouts, reflect systemic inefficiencies that erode patient trust and motivation, these findings are similar to what is reported in South Africa and in a narrative review conducted by systematically searching electronic databases (39, 40). Additionally, patients’ perceptions of being well and symptom-free, coupled with physical limitations, diminish their perceived necessity for ongoing care, consistent with findings from studies in the UK, USA, and Spain on low risk perception in asymptomatic hypertension (41-43).

## Study limitation

This study faced challenges related to missing data as it involved retrospective review of patient files, some variables such as education level was not documented in patient records and thus could not be analyzed, potentially omitting important confounders. The use of consecutive sampling in the quantitative phase may have introduced selection bias, limiting the generalizability of the findings to the broader patient population. In the qualitative component, reliance on participants’ recall of past clinic experiences may have introduced recall bias, as some may have forgotten, misremembered, or provided socially desirable responses.

## Conclusions

This study found a substantial attrition rate of 56.8%, with the highest number of patients lost to follow-up occurring in 2020, coinciding with the peak of the COVID-19 pandemic. Various factors were associated with time to attrition, including age, male sex, residing outside the capital city, and last visit blood pressure measurements. Furthermore, loss to follow-up was driven by structural and contextual barriers, health system challenges, and illness perceptions and health-related limitations. These included financial hardship, long distances, overcrowding, provider attitudes, and perceived lack of treatment benefit.

These findings imply that retention rates are considerably below the Centers for Disease Control and Prevention’s 80% retention target, highlighting the need to implement targeted strategies to enhance patient retention, particularly among high risk groups. Strengthening tracking systems, improving access to healthcare, and minimizing the effects of external disruptions such as pandemics may support sustained patient engagement.

## Supporting information

IRB ethical approval

Informed_consent_qualitative

Consent_waiver_quantitative

Anonymised dataset

## Data Availability

All relevant data are within the manuscript and its Supporting Information files

## Acknowledgements

We are grateful to the study participants and the clinic staff at Mulago hospital hypertension clinic for the support rendered to make this study a success.

## Financial Disclosure

There was no funding for this study.

## Competing interests

There were no competing interests for all authors. No organization played any role in the study design, data collection and analysis, decision to publish, or preparation of the manuscript.

## Availability of data and Materials

All the necessary data and materials underlying the findings described have been provided as part of this manuscript.

## Author Contributions

Conceptualization: NN, JM & JNK

Formal analysis: NN, BB, PM, MO &SL

Data curation: NN, MO

Funding acquisition: NN

Methodology: NN, JNK, BB, MB, MO & SL

Project administration: NN

Resources: NN

Supervision: JNK, MKM, PM &NN

Writing original draft: NN, JNK, MKM

Writing review & editing: JNK, MKM, PM, DB, SL, BB, MO, JM, MB & NN

## Supporting information

‘’Cover letter’’

‘’Fig 1’’

‘’Fig 2’’

‘’Fig 3’’

‘’IRB approval letter’’

**‘’** IRB approval of consent waiver’’

‘’Anonymized quantitative dataset’’

‘’Informed consent’’

## Notes

### Competing Interest Statement

The authors have declared no competing interest.

### Funding Statement

The author(s) received no specific funding for this work.

### Author Declarations

School of Medicine Research and Ethics Committee (SOMREC) of Makerere University, College of Health Sciences gave ethical approval for this work.

## References

1. Allen LN, Pullar J, Wickramasinghe KK, Williams J, Roberts N, Mikkelsen B, et al. Evaluation of research on interventions aligned to WHO ‘Best Buys’ for NCDs in low-income and lower-middle-income countries: a systematic review from 1990 to 2015. 2018;3(1):e000535.

2. WHO. Hypertension 2023 [Available from: https://www.who.int/news-room/fact-sheets/detail/hypertension#:~:text=Key%20facts%201%20An%20estimated%201.28%20billion%20adults,with%20hypertension%20have%20it%20under%20control.%20More%20items.

3. Zhou B, Carrillo-Larco RM, Danaei G, Riley LM, Paciorek CJ, Stevens GA, et al. Worldwide trends in hypertension prevalence and progress in treatment and control from 1990 to 2019: a pooled analysis of 1201 population-representative studies with 104 million participants. The lancet. 2021;398(10304):957–80.

4. Guwatudde D, Mutungi G, Wesonga R, Kajjura R, Kasule H, Muwonge J, et al. The epidemiology of hypertension in Uganda: findings from the national non-communicable diseases risk factor survey. 2015;10(9):e0138991.

5. Guwatudde D, Mutungi G, Wesonga R, Kajjura R, Kasule H, Muwonge J, et al. The epidemiology of hypertension in Uganda: findings from the national non-communicable diseases risk factor survey. PloS one. 2015;10(9):e0138991.

6. Kubiak RW, Sveum EM, Faustin Z, Muwonge T, Zaidi HA, Kambugu A, et al. Prevalence and risk factors for hypertension and diabetes among those screened in a refugee settlement in Uganda. 2021;15:1–8.

7. Frieden TR, Jaffe MGJTJoCH. Saving 100 million lives by improving global treatment of hypertension and reducing cardiovascular disease risk factors. 2018;20(2):208.

8. Ye J, Orji IA, Baldridge AS, Ojo TM, Shedul G, Ugwuneji EN, et al. Characteristics and patterns of retention in hypertension care in primary care settings from the hypertension treatment in Nigeria Program. 2022;5(9):e2230025-e.

9. Dal Canto E, Ceriello A, Rydén L, Ferrini M, Hansen TB, Schnell O, et al. Diabetes as a cardiovascular risk factor: An overview of global trends of macro and micro vascular complications. 2019;26(2_suppl):25-32.

10. Nikpour Hernandez N, Ismail S, Heang H, van Pelt M, Witham MD, Davies JIJHP, et al. An innovative model for management of cardiovascular disease risk factors in the low resource setting of Cambodia. 2021;36(4):397–406.

11. Ma C, Chen S, Zhou Y, Huang CJANR. Treatment adherence of Chinese patients with hypertension: A longitudinal study. 2013;26(4):225–31.

12. Degoulet P, Menard J, Vu H, Golmard J, Devries C, Chatellier G, et al. Factors predictive of attendance at clinic and blood pressure control in hypertensive patients. Br Med J (Clin Res Ed). 1983;287(6385):88-93.

13. Nikpour Hernandez N, Ismail S, Heang H, van Pelt M, Witham MD, Davies JI. An innovative model for management of cardiovascular disease risk factors in the low resource setting of Cambodia. Health Policy and Planning. 2021;36(4):397–406.

14. Barreto MdS, Reiners AAO, Marcon SS. Knowledge about hypertension and factors associated with the non-adherence to drug therapy. Revista latino-americana de enfermagem. 2014;22(03):491–8.

15. Flaus-Furmaniuk A, Fianu A, Lenclume V, Chirpaz E, Balcou-Debussche M, Debussche X, et al. Attrition and social vulnerability during 2-year-long structured care in type 2 diabetes, the ERMIES randomized controlled trial. BMC Endocrine Disorders. 2022;22(1):314.

16. Gebre TE, Bekele AJIJArp. Attrition rate and predictors among adult patients receiving antiretroviral therapy in Adama Hospital Medical College, Central Ethiopia: a retrospective cohort study design, 2006–2017. 2019;8(3):2456-9992.

17. Alizadeh F, Mfitumuhoza G, Stephens J, Habimaana C, Myles K, Baganizi M, et al. Identifying and reengaging patients lost to follow-up in rural Africa: the “horizontal” hospital-based approach in Uganda. 2019;7(1):103–15.

18. Tashakkori A, Creswell JWJJommr. The new era of mixed methods. Sage Publications; 2007. p. 3-7.

19. Israel GD. Determining sample size. 1992.

20. Meena J, Raghav P, Rustagi N. LBOS 03-06 anti hypertensive treatment compliance and adverse effect profile among hypertension clinic attendees in Jodhpur, India. Journal of Hypertension. 2016;34:e552.

21. Given CW, Given BA, Coyle BW. Prediction of patient attrition from experimental behavioral interventions. Nursing research. 1985;34(5):293–8.

22. John W, Creswell P, Cheryl N. Qualitative inquiry and research designinternational student edition: Choosing Among… Five Approaches: SAGE PUBLICATIONS Incorporated; 2024.

23. Maguire M, Delahunt BJAIJoHE. Doing a thematic analysis: A practical, step-by-step guide for learning and teaching scholars. 2017;9(3).

24. Engelland AL, Alderman MH, Powell HBJAJoPH. Blood pressure control in private practice: a case report. 1979;69(1):25–9.

25. Ramsay JA, McKENZIE JK, Fish DGJAJoPH. Physicians and nurse practitioners: do they provide equivalent health care? 1982;72(1):55–7.

26. Meena J, Raghav P, Rustagi NJJoH. LBOS 03-06 anti hypertensive treatment compliance and adverse effect profile among hypertension clinic attendees in Jodhpur, India. 2016;34:e552.

27. Kassavou A, Mirzaei V, Shpendi S, Brimicombe J, Chauhan J, Bhattacharya D, et al. The feasibility of the PAM intervention to support treatment-adherence in people with hypertension in primary care: a randomised clinical controlled trial. 2021;11(1):8897.

28. Baldé A, Lièvre L, Maiga AI, Diallo F, Maiga IA, Costagliola D, et al. Risk factors for loss to followCup, transfer or death among people living with HIV on their first antiretroviral therapy regimen in Mali. HIV medicine. 2019;20(1):47–53.

29. Degoulet P, Menard J, Vu H, Golmard J, Devries C, Chatellier G, et al. Factors predictive of attendance at clinic and blood pressure control in hypertensive patients. 1983;287(6385):88-93.

30. Hernandez NN, Ismail S, Heang H, van Pelt M, Witham MD, Davies JIJHP, et al. An innovative model for management of cardiovascular disease risk factors in the low resource setting of Cambodia. 2021;36(4):397.

31. Given CW, Given BA, Coyle BWJNr. Prediction of patient attrition from experimental behavioral interventions. 1985;34(5):293–8.

32. Zhu J, Yousuf MA, Yang W, Zhu Q, Shen Z, Lan G, et al. Mortality and attrition rates within the first year of antiretroviral therapy initiation among people living with HIV in Guangxi, China: an observational cohort study. 2021;2021(1):6657112.

33. Flaus-Furmaniuk A, Fianu A, Lenclume V, Chirpaz E, Balcou-Debussche M, Debussche X, et al. Attrition and social vulnerability during 2-year-long structured care in type 2 diabetes, the ERMIES randomized controlled trial. 2022;22(1):314.

34. Zemariam AB, Abebe GK, Alamaw AWJSR. Incidence and predictors of attrition among human immunodeficiency virus infected children on antiretroviral therapy in Amhara comprehensive specialized hospitals, Northwest Ethiopia, 2022: a retrospective cohort study. 2024;14(1):4366.

35. Pasquet A, Messou E, Gabillard D, Minga A, Depoulosky A, Deuffic-Burban S, et al. Impact of drug stock-outs on death and retention to care among HIV-infected patients on combination antiretroviral therapy in Abidjan, Côte d’Ivoire. 2010;5(10):e13414.

36. GhanbariCJahromi M, Kharazmi E, Bastani P, Shams M, Marzaleh MA, Amin Bahrami M. Factors disrupting the continuity of care for patients with chronic disease during the pandemics: A systematic review. Health science reports. 2024;7(2):e1881.

37. Ljungholm L, Edin-Liljegren A, Ekstedt M, Klinga C. What is needed for continuity of care and how can we achieve it? –Perceptions among multiprofessionals on the chronic care trajectory. BMC health services research. 2022;22(1):686.

38. Jeon Y-H, Essue B, Jan S, Wells R, Whitworth JA. Economic hardship associated with managing chronic illness: a qualitative inquiry. BMC health services research. 2009;9:1–11.

39. Tana VV. Experiences of chronic patients about long waiting time at a community health care centre in the Western Cape. 2013.

40. Anderson J. Barriers to Achieving Continuity of Care in Primary Health Care: A Narrative Review of Challenges and Solutions. 2020.

41. Ross S, Walker A, MacLeod MJ. Patient compliance in hypertension: role of illness perceptions and treatment beliefs. Journal of human hypertension. 2004;18(9):607–13.

42. Williams GH. Assessing patient wellness: new perspectives on quality of life and compliance. American journal of hypertension. 1998;11(S8):186S–91S.

43. Chaudhry A. A perception-based approach to healthcare: Understanding perspectives for improved outcomeS. 2024.

