## Supplementary material for "Attrition and associated factors among patients on chronic antihypertensive therapy at Mulago hospital, Uganda: A mixed method study": IRB ethical approval

**Appendix I: IRB approval letter**

MAKERERE UNIVERSITY

P.O. Box 7072 Kampala, Uganda


COLLEGE OF HEALTH SCIENCES
SCHOOL OF MEDICINE

RESEARCH ETHICS COMMITTEE

**30/05/2023**

To: Nathan Ntenkaire

0786051858

**Type:** Initial Review

**Re: Mak-SOMREC-2023-584: Attrition and associated risk factors among patients on chronic antihypertensive therapy at Mulago hospital**

I am pleased to inform you that at the **146** convened meeting on **20/04/2023**, the MAK School of Medicine REC (Mak-SOMREC), committee meeting voted to approve the above referenced application.
Approval of the research is for the period of **30/05/2023** to **30/05/2024.**

As Principal Investigator of the research, you are responsible for fulfilling the following requirements of approval:

1. All co-investigators must be kept informed of the status of the research.
2. Changes, amendments, and addenda to the protocol or the consent form must be submitted to the REC for re-review and approval **prior** to the activation of the changes.
3. Reports of unanticipated problems involving risks to participants or any new information which could change the risk benefit: ratio must be submitted to the REC.
4. Only approved consent forms are to be used in the enrollment of participants. All consent forms signed by participants and/or witnesses should be retained on file. The REC may conduct audits of all study records, and consent documentation may be part of such audits.
5. Continuing review application must be submitted to the REC **eight weeks** prior to the expiration date of **30/05/2024** in order to continue the study beyond the approved period. Failure to submit a continuing review application in a timely fashion may result in suspension or termination of the study.
6. The REC application number assigned to the research should be cited in any correspondence with the REC of record.
7. You are required to register the research protocol with the Uganda National Council for Science and Technology (UNCST) for final clearance to undertake the study in Uganda.

The following is the list of all documents approved in this application by MAK School of Medicine REC (Mak-SOMREC):

| No. | Document Title | Language | Version Number | Version Date |
|---|---|---|---|---|
| 1 | Protocol | English | 4.0 | 2023-05-17 |
| 2 | Informed consent form for the recruitment of research participants | Luganda | 1.0 | 2023-05-16 |
| 3 | Informed consent form for the recruitment of research participants | English | 1.0 | 2023-05-16 |
| 4 | COVID-19 & EBOLA risk management plan for the study | English | 1.0 | 2023--13 |
| 5 | Data collection tools | Luganda | 1.0 | 2023-03-10 |
| 6 | Data collection tools | English | 1.0 | 2023-03-10 |
| 7 | Data collection tools | English | 1.0 | 2023-03-10 |
| 8 | Application for waiver of informed consent if applicable to your study | English | 1.0 | 2023-03-10 |

Yours Sincerely

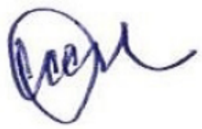

Prof. Ponsiano Ocama
For: MAK School of Medicine REC (Mak-SOMREC)
