## Supplementary material for "Attrition and associated factors among patients on chronic antihypertensive therapy at Mulago hospital, Uganda: A mixed method study": Informed_consent_qualitative

### Informed consent form (English)

**Principal investigator**: Nathan Ntenkaire

**Date**: 3/17/2023

**Background for the study:** Loss to follow up among patients on chronic treatment of hypertension is a significant problem that can lead to serious health consequences, including uncontrolled blood pressure levels, cardiovascular diseases, stroke, sexual dysfunction, sudden loss of kidney functions, and blindness. Multiple risk factors may contribute to this issue, and it is crucial for healthcare providers to be mindful of these factors to prevent treatment discontinuation and ensure optimal patient outcomes. The study will involve reviewing patients' medical records in the quantitative phase and in-depth phone interviews shall be conducted in the qualitative phase.

**Rationale for the study:** The study's outcomes will aid policymakers in determining the most efficient approach to addressing attrition-related concerns and provide insights into innovative methods for reaching and retaining patients on antihypertensive therapy. Additionally, by identifying specific patient groups at greater risk of loss to follow up, targeted care and support can be offered. The findings will also assist Mulago hospital administration in developing strategies to decrease attrition and improve patient retention.

**Purpose:** To determine the underlying reasons for loss to follow up among patients who were initiated on hypertension treatment between January 2020 and December 2022 at Mulago hospital hypertension clinic.

**Procedures:** If you decide to take part in this study, the research assistant (registered nurse) will ask you a few questions regarding the underlying reasons for your loss to follow up and any knowledge about hypertension and its treatment. This process will take not more than 30 minutes of your time and the interview shall be audio recorded.

MAKERERE UNIVERSITY
SCHOOL OF MEDICINE
INITIAL APPROVAL
VALID UNTIL
30 MAY 2024
RESEARCH & ETHICS COMMITTEE
P.O. BOX 7072,KAMPALA

**Questions about participants rights:** By agreeing to participate in the study, you do NOT waive any of your legal rights. If you have any questions about the study, please contact the PI **Nathan Ntenkaire Tel: 0786051858.**

**Questions:** If you have any questions or concerns regarding your rights as a study participant and would like to talk to someone other than the researcher(s), please feel free to contact **Prof. Ponsiano Ocama**, chairman of Makerere University School of Medicine Research and Ethics Committee (SOMREC), on **0772421190.**

**Statement of voluntariness:** By agreeing to take part, I do not waive my rights but show that I voluntarily decide to join this study and that I have a right to withdraw my participation at any time without any penalty.

**Dissemination of results:** Copies of the research study shall be submitted to the Clinical Epidemiology Unit, Albert cook library and Directorate of Research and Graduate training. This research work shall be published in a peer reviewed scientific journal and the research findings shall be discussed with the MH-HTN clinic staff

**Certificate of Consent:**

The consent form has been read to me. Have understood the procedures, risks and benefits of participating in this study. I understand that my decision to participate in this study is completely voluntary and that I have a right to withdraw at any time without any penalty. I understand that by signing this form, I do not waive any of my legal rights but merely indicate that I have been informed about the research study in which I am voluntarily agreeing to participate. I have had the opportunity to ask questions and had the questions answered satisfactorily. I voluntarily consent to participate in this study.

Name of participant.......................................................Signature/thumbprint.................

Date..............................

Name of witness................................ Signature..............................

Date..............................

MAKERERE UNIVERSITY
SCHOOL OF MEDICINE
INITIAL APPROVAL
VALID UNTIL
30 MAY 2024
RESEARCH & ETHICS COMMITTEE
P.O. BOX 7072,KAMPALA
