## Supplementary material for "Attrition and associated factors among patients on chronic antihypertensive therapy at Mulago hospital, Uganda: A mixed method study": Consent_waiver_quantitative

**Appendix J: IRB approval of consent waiver**

MAKERERE UNIVERSITY
P.O. Box 7072 Kampala, Uganda

**COLLEGE OF HEALTH SCIENCES**
**SCHOOL OF MEDICINE**

RESEARCH ETHICS COMMITTEE

May 30, 2023

Mr. Nathan Ntenkaire
Clinical Epidemiology Unit

Dear Mr. Ntenkaire,

**RE: APPROVAL OF CONSENT WAIVER**

In your letter dated 10th March 2023, you requested the committee to waive off consent for the study entitled **"Attrition and associated risk factors among patients on chronic antihypertensive therapy at Mulago hospital"**. The study involves review of medical records of patients diagnosed with hypertension and initiated on AHT at the Mulago hospital hypertension clinic between January 2020 and December 2022.

On behalf of the committee, I am glad to inform you that the committee has granted waiver of the informed consent process for the retrospective part of the study.

Yours sincerely,

Dr. Aloysius Gonzaga Mubuuke
**Vice Chairperson School of Medicine Research and Ethics Committee**
